# Genetic Factors Associated with Prostate Cancer Conversion from Active Surveillance to Treatment

**DOI:** 10.1101/2021.08.30.21262305

**Authors:** Yu Jiang, Travis J. Meyers, Adaeze A. Emeka, Lauren Folgosa Cooley, Phillip R. Cooper, Nicola Lancki, Irene Helenowski, Linda Kachuri, Daniel W. Lin, Janet L. Stanford, Lisa F. Newcomb, Suzanne Kolb, Antonio Finelli, Neil E. Fleshner, Maria Komisarenko, James A. Eastham, Behfar Ehdaie, Nicole Benfante, Christopher J. Logothetis, Justin R. Gregg, Cherie A. Perez, Sergio Garza, Jeri Kim, Leonard S. Marks, Merdie Delfin, Danielle Barsa, Danny Vesprini, Laurence H. Klotz, Andrew Loblaw, Alexandre Mamedov, S. Larry Goldenberg, Celestia S. Higano, Maria Spillane, Eugenia Wu, H. Ballentine Carter, Christian P. Pavlovich, Mufaddal Mamawala, Tricia Landis, Peter R. Carroll, June M. Chan, Matthew R. Cooperberg, Janet E. Cowan, Todd M. Morgan, Javed Siddiqui, Rabia Martin, Eric A. Klein, Karen Brittain, Paige Gotwald, Daniel A. Barocas, Jeremiah R. Dallmer, Jennifer B. Gordetsky, Pam Steele, Shilajit D. Kundu, Jazmine Stockdale, Monique J. Roobol, Lionne D.F. Venderbos, Martin G. Sanda, Rebecca Arnold, Dattatraya Patil, Christopher P. Evans, Marc A. Dall’Era, Anjali Vij, Anthony J. Costello, Ken Chow, Niall M. Corcoran, Soroush Rais-Bahrami, Courtney Phares, Douglas S. Scherr, Thomas Flynn, R. Jeffrey Karnes, Michael Koch, Courtney Rose Dhondt, Joel B. Nelson, Dawn McBride, Michael S. Cookson, Kelly L. Stratton, Stephen Farriester, Erin Hemken, Walter M. Stadler, Tuula Pera, Deimante Banionyte, Fernando J. Bianco, Isabel H. Lopez, Stacy Loeb, Samir S. Taneja, Nataliya Byrne, Christopher L. Amling, Ann Martinez, Luc Boileau, Franklin D. Gaylis, Jacqueline Petkewicz, Nicholas Kirwen, Brian T. Helfand, Jianfeng Xu, Denise M. Scholtens, William J. Catalona, John S. Witte

**Author notes:** These authors contributed equally to the first authorship (Yu Jiang, Travis J. Meyers). These authors contributed equally to the senior authorship (William J. Catalona, John S. Witte). Correspondence to John S. Witte.

## Abstract

Men diagnosed with low-risk prostate cancer (PC) are increasingly electing active surveillance (AS) as their initial management strategy. While this may reduce the side effects of treatment for prostate cancer, many men on AS eventually convert to active treatment. PC is one of the most heritable cancers, and genetic factors that predispose to aggressive tumors may help distinguish men who are more likely to discontinue AS. To investigate this, we undertook a multi-institutional genome-wide association study (GWAS) of 6,361 PC patients who initially elected AS and were followed over time for the potential outcome of conversion from AS to active treatment. In the GWAS we detected 18 single nucleotide polymorphisms (SNPs) associated with conversion, 15 of which were not previously associated with PC risk. We found two genes associated with conversion (*MAST3*, p = 6.9×10^-7^ and *GAB2*, p = 2.0×10^-6^). Moreover, increasing values of a previously validated 269-SNP genetic risk score (GRS) for PC was positively associated with conversion (e.g., comparing the highest to the two middle deciles gave a hazard ratio [HR] = 1.13; 95% Confidence Interval [CI]= 0.94-1.36); whereas, decreasing values of a 36-variant GRS for prostate-specific antigen (PSA) levels were positively associated with conversion (e.g., comparing the lowest to the two middle deciles gave a HR = 1.25; 95% CI, 1.04-1.50). These results suggest that germline genetics may help inform and individualize the decision of AS—or the intensity of monitoring on AS—*versus* treatment for the initial management of patients with low-risk PC.

## Introduction

Active surveillance (AS) is now more widely implemented as an initial management strategy for many men with lower-risk prostate cancer (PC [MIM: 176807])^1^. PC that is unlikely to invade surrounding tissue or metastasize according to characteristics at diagnosis is considered “low-risk” or “favorable-intermediate risk”^2^. Recent work in the United States Veterans Administration (VA) Health Care System^3, 4^ and in Sweden^5^ indicates that a majority of men with low-risk PC are being managed with AS. Determining which patients most benefit from early active treatment *versus* AS, however, and how intensive the surveillance protocol should be, remains a challenge.

A major drawback of AS for low-risk PC is the possibility of misclassifying patients with life-threatening disease. In fact, over a 10-year follow-up period, 20-40% of men initially managed with AS later have more aggressive cancer^6^. While the impact of delayed treatment is unknown, up to 50% of men in one AS series of studies experienced biochemical recurrence after active treatment^7^. These uncertainties and challenges in accurately discriminating between indolent and aggressive PC may prompt men to err on the side of early treatment resulting in unnecessary side effects and worse health-related quality of life, or conversely result in delays in therapy for men who are likely to benefit from it. Many men have such low-risk disease that they do not need the biopsies or scans with the frequency with which they have typically been performed. Recent work suggests that the likelihood of risk reclassification of an individual patient’s disease might be predicted for at least four years of AS^8^. Thus, it may be possible reduce the intensity of surveillance for many men with the lowest-risk tumors.

A key outstanding question is how to best distinguish among low- and high-risk tumors for AS decisions. Promising recent developments for enhancing clinical risk assessment include multi-parametric MRI with targeted prostate biopsy and tissue-based genomic testing^9, 10^. Another potentially valuable approach is incorporating germline genetic information for PC via a polygenic risk score^11^. PC is one of the most heritable of common cancers, with germline genetic factors accounting for over 40% of the variability in this disease^12–15^. We and others have identified from genome-wide association studies (GWAS) 269 common germline genetic variants associated with PC susceptibility that explain a substantial proportion of disease heritability^16–43^. Combining these PC risk variants together into a genetic risk score (GRS) may provide a more discriminatory biomarker not only for PC risk, but also potentially for predicting conversion from AS to treatment^44–47^. Moreover, we recently have discovered genetic variants that explain variability in PSA levels^48^. Since PSA is a critical component to monitoring men undergoing AS, incorporating this information may also help to identify ideal AS candidates.

To evaluate the potential value of incorporating germline genetic information into the shared decision-making process for AS, we present findings from a large, multi-institutional GWAS of men diagnosed with PC enrolled in an AS program. We report novel variants and genes, and genetic risk scores associated with conversion from AS to treatment.

## Material and Methods

### Participants

The primary study participants came from 28 institutions in the US, Canada, the Netherlands, and Australia. We recently reported on the clinicopathological characteristics of conversion to treatment in this population^49^. The initial study population included 7,279 men diagnosed with PC between 1991 and 2018 who elected AS for their initial management. We also included an additional 593 AS patients from the University of Texas MD Anderson Cancer Center as replication samples, described below. Patients’ blood or tissue samples were collected to conduct germline genetic analyses. The AS protocols varied among participating institutions, reflecting real-world practice patterns^50–53^, and we did not impose strict inclusion/exclusion criteria based on the AS protocol. Patient demographic and clinical variables were collected and managed using the Research Electronic Data Capture (REDCap) software^54, 55^. Men without data on the duration of AS (235, 3.2%) and those managed with AS for less than six months (269, 3.7%) were excluded, leaving 6,775 men for potential inclusion in the GWAS. This study was approved by the institutional review board at each institution, all participants provided written informed consent, and all participating institutions signed a material transfer and data use agreement.

### Clinical and Demographic Factors

We collected PC characteristics at diagnosis, including the age at diagnosis, Gleason grade group (GG), PSA level, clinical tumor stage (cT), and the number of cancerous biopsy cores at diagnosis. Grade groups correspond to the following Gleason scores (GS): GG1 ∼ GS ≤6; GG2 ∼ GS 3+4; GG3 ∼ GS 4+3; GG4 ∼ GS 8; GS 5 ∼ GS 9 or 10^56^. Study participants were classified into three risk groups (low-, intermediate-, and high-risk) based on our modification of guidelines from the National Comprehensive Cancer Network (NCCN) and the American Urological Association (AUA). We did not strictly follow these guidelines because we were unable to distinguish between cT2a, cT2b, and cT2c, and we did not have data on PSA density (serum PSA concentration divided by prostate volume). Therefore, *low-risk* patients met the following criteria: Gleason grade group 1 (GG1) (Gleason score 3+3), PSA <10 ng/mL, clinical-stage cT1, and ≤3 positive biopsy cores. *Intermediate-risk* patients had any of the following without any high-risk or high-volume criteria: GG2 (Gleason 3+4), PSA 10-20 ng/mL, or stage cT2. *High-risk* patients had any of the following: ≥ GG3 (≥ Gleason 4+3), PSA ≥20 ng/mL, stage ≥cT3, or ≥4 positive biopsy cores of any GG.

Conversion occurred when a patient received treatment following AS. The reason for withdrawing from AS to begin treatment was reported as due to “upgrading”, “upstaging”, “PSA progression”, “anxiety”, and/or “other” reasons. Note that in our survival analysis (below), individuals who converted due to anxiety were censored and do not contribute events in our analysis. We used the ADMIXTURE software program to infer genetic ancestry from uncorrelated SNPs, according to major reference populations in the 1000 Genomes Project (European, African, East/South Asian combined, and Admixed American)^57^.

### Genotyping and Imputation

In total, 6,324 participants were genotyped on the Illumina Infinium Multi-Ethnic Global Array (MEGA), including custom content, at the NIH Center for Inherited Disease Research (CIDR) at Johns Hopkins University. Genotypes were called using GenomeStudio version 2011.1, Genotyping Module version 1.9.4, and GenTrain Version 1.0. The full array with custom content consisted of 1,760,143 SNPs.

After genotyping, the median SNP call rate was 99.94%, and the error rate estimated from 122 pairs of planned study duplicates was 1.3×10^-6^. Samples and variants were excluded if they had a sample or genotyping call rate < 98%. We limited our analyses to variants with a minor allele frequency (MAF) ≥ 1%. Variants were screened for deviations from Hardy-Weinberg equilibrium with a filter threshold of p=6.5×10^-4^. A total of 856,077 genotyped SNPs remained after these quality control (QC) steps. Unmeasured genetic variants were imputed using the Trans-Omics for Precision Medicine (TOPMed) Imputation Server, with 97,256 reference samples and 308,107,085 SNPs. Variants with imputation quality (INFO) score < 0.3 were excluded, leaving a total of 22,691,641 SNPs successfully imputed. After QC steps, a total of 5,936 samples genotyped at CIDR remained for inclusion in the GWAS.

Furthermore, we included in our analysis an additional 593 AS patients from MD Anderson previously genotyped on the Illumina Infinium OncoArray-500K BeadChip Array. This array was primarily developed to study cancer predisposition and risk. Genotypes were called using GenomeStudio version 2011.1. The full array consisted of 500,000 SNPs. Genotype QC procedures and imputation for the PRACTICAL OncoArray have been described previously^15^. Briefly, imputation was performed without pre-phasing with SHAPEIT2 based on the 1000 Genomes Phase 3 release reference panel. In total, 21,299,194 SNPs were successfully imputed, and 10,109,977 variants with MAF ≥ 1% on autosomal chromosomes 1-22 and sex chromosome X.

### GWAS of Conversion from AS to Treatment

The variants with MAF ≥ 1% on autosomal chromosomes 1-22 and sex chromosome X were tested for their association with time to conversion from AS to treatment among the 5,222 men of European genetic ancestry genotyped by CIDR. Patients who converted due to anxiety were censored because the event of interest was converting due to a change in the cancer clinical characteristics. Per-allele hazard ratios (HRs), 95% confidence intervals (CIs), and corresponding p-values were calculated from Cox proportional hazards models. HRs were adjusted for age at diagnosis and the first 10 genetic principal components to address potential population stratification or cryptic relatedness. Adjusted HRs were calculated using the gwasurvivr package in R^58^. For any variants associated with conversion, we examined the Cox models’ proportional hazards assumption.

Following the GWAS discovery phase, the potential associations were tested for replication in an independent GWAS among 1,139 men also genotyped by CIDR (but of non-European ancestry) and the 425 MD Anderson samples of European genetic ancestry (excluding other ancestries). Again, variants with MAF ≥ 1% on autosomal chromosomes and sex chromosome X were tested for their association with conversion within major ancestral populations (i.e., European, African, Asian, and Admixed American). For the MD Anderson patients, 9,962,324 variants were tested in a Cox proportional hazards model adjusted for age at PC diagnosis and ancestry principal components.

Results from the GWAS were combined with a fixed-effects inverse-variance-weighted meta-analysis using METAL^59^. All statistical tests were two-sided. Marginal P values less than 5×10^-8^ were considered statistically significant. We defined a locus as the 1 megabase (Mb) region surrounding the sentinel variant (500 kilobase pairs flanking each side). To identify independently associated variants, within each 1 Mb region we performed clumping on the association results using PLINK v1.9 using a linkage disequilibrium threshold r^2^ < 0.5).

### Transcriptome-Wide Association Study of Conversion from AS to Treatment

To identify additional genes associated with time to conversion, we conducted a transcriptome-wide association analysis (TWAS), which models genetically imputed transcript levels and has a lower multiple testing burden compared to single-variant analysis. We applied the MetaXcan analytic pipeline to our GWAS summary statistics and associated genetically predicted expression of approximately 22,000 genes across 49 issue reference datasets from GTEx (version 8)^60^. Tissue-specific associations were aggregated using S-MultiXcan to obtain cross-tissue p-values for each gene^60^. Associations were considered statistically significant at the Bonferroni-corrected alpha level of 2.2×10^-6^ (i.e., 0.05/22,535 genes).

### Genetic Risk Scores

Genetic risk scores (GRS) were constructed by summing variant-specific weighted allelic dosages for the samples genotyped by CIDR. The initial GRS included the 269 PC risk variants reported in the largest trans-ancestry GWAS meta-analysis of PC^43^. Specifically, for patient *i*, 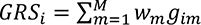, where *g_im_* is the genotype dosage for patient *i*, and variant *m*, and *w_m_* is the variant weight on the log odds ratio scale from the published literature (i.e., the meta-analysis for the GRS_PC_). *M* is the total number of variants included in the GRS (M = 269 for the GRS_PC_). A second GRS was developed for the genetic basis of serum PSA levels. This GRS_PSA_ included 36 variants and their corresponding weights from a GWAS of PSA levels among cancer-free men.^48^ The associations between these GRS and conversion were estimated using multivariable Cox proportional hazards models. Minimally adjusted Cox models included age and the first 10 genetic principal components. Fully adjusted Cox models also included Gleason grade group (GG1, GG2, or ≥ GG3); PSA concentration (ng/mL); clinical stage (cT1, cT2, or cT3/cT4); and number of positive biopsy cores (1-2, 3, or ≥ 4). The GRS was modeled as a categorical variable according to deciles of the distribution, and HRs for each decile were estimated with the 40-60th percentile group as the reference.

### Clinical Utility of the GRS

The potential utility of the GRS was evaluated by comparing how the top and bottom deciles of the GRS distribution modified conversion rates within the three PC clinicopathological risk categories (i.e., low-, intermediate-, and high-). For the top and bottom GRS deciles (top 10^th^ percentile and bottom 10^th^ percentile, respectively) we plotted Kaplan–Meier curves of conversion within each PC clinicopathological risk category and tested the difference between each pair of curves with the log-rank test.

To evaluate the overall discriminative capacity of the GRS (i.e., not just the decile tails), we calculated the area under the ROC Curve (AUC) in the discovery sample using regression models of time to conversion. We used Chambless and Diao’s estimator of cumulative AUC for right-censored time-to-event data, which is a summary measure given by the integral of AUC on [0, max(times)] weighted by the estimated probability density of the time-to-event outcome^61^. A baseline AUC was calculated for the model that included age and the first 10 principal components. This model was then expanded to further include PC clinical characteristics listed above for the multivariable Cox model, followed by GRS_PC_ and GRS_PSA_ (individually and together).

## Results

### Study Population

Details of the discovery and replication samples that met inclusion criteria are presented in **Table S1**. Clinically, most men had low-risk PC (3,639, 70%) and/or features of low-risk, low-volume disease: GG1 (4,819, 92%), 1-2 positive biopsy cores (4,113, 79%), and a median PSA at diagnosis of 5 ng/mL. Using genetic information to infer ancestry, 88% were classified as European ancestry. The demographic and clinicopathological characteristics of the replication samples had a similar pattern as the discovery samples, except that the proportion of high-risk PC was higher for men of Asian genetic ancestry (n = 43; 18%) than of European ancestry (n= 599; 11%); **Table S1**). Baseline characteristics were missing for the following proportion of study participants: age at diagnosis (<0.1%), GG group (<0.1%), PSA concentration (3.3%), clinical tumor stage (6.9%), number of positive biopsy cores (2.5%), and risk-group classification (<0.1%).

### Genome-Wide Association Study of Conversion from AS to Treatment

Our approach to the GWAS discovery, replication, and meta-analysis is outlined in Figure 1. The median follow-up time for patients in this multicenter study was 6.7 years. Of the 2,260 patients who converted from AS to treatment, 126 (5.6%) converted due to anxiety and were censored in our time-to-event analyses. Our primary discovery GWAS yielded 14 independent lead SNPs (i.e., p-value < 5 x 10^-8^ at each locus of size 1Mb) (Figure 2a). We replicated 1 of the signals at a p-value level less than 0.05/14 (≈ 0.0036) in the replication meta-analysis. In the combined meta-analysis of discovery and replication GWAS, we detected four additional SNPs independently associated with conversion to treatment (Figure 2b). Q-Q plots for the discovery GWAS and the combined meta-analysis did not suggest inflation of test statistics due to systematic bias such as population substructure (genomic inflation factor =1 and 1.02, respectively; **Figure S1**).

**Figure 1.**
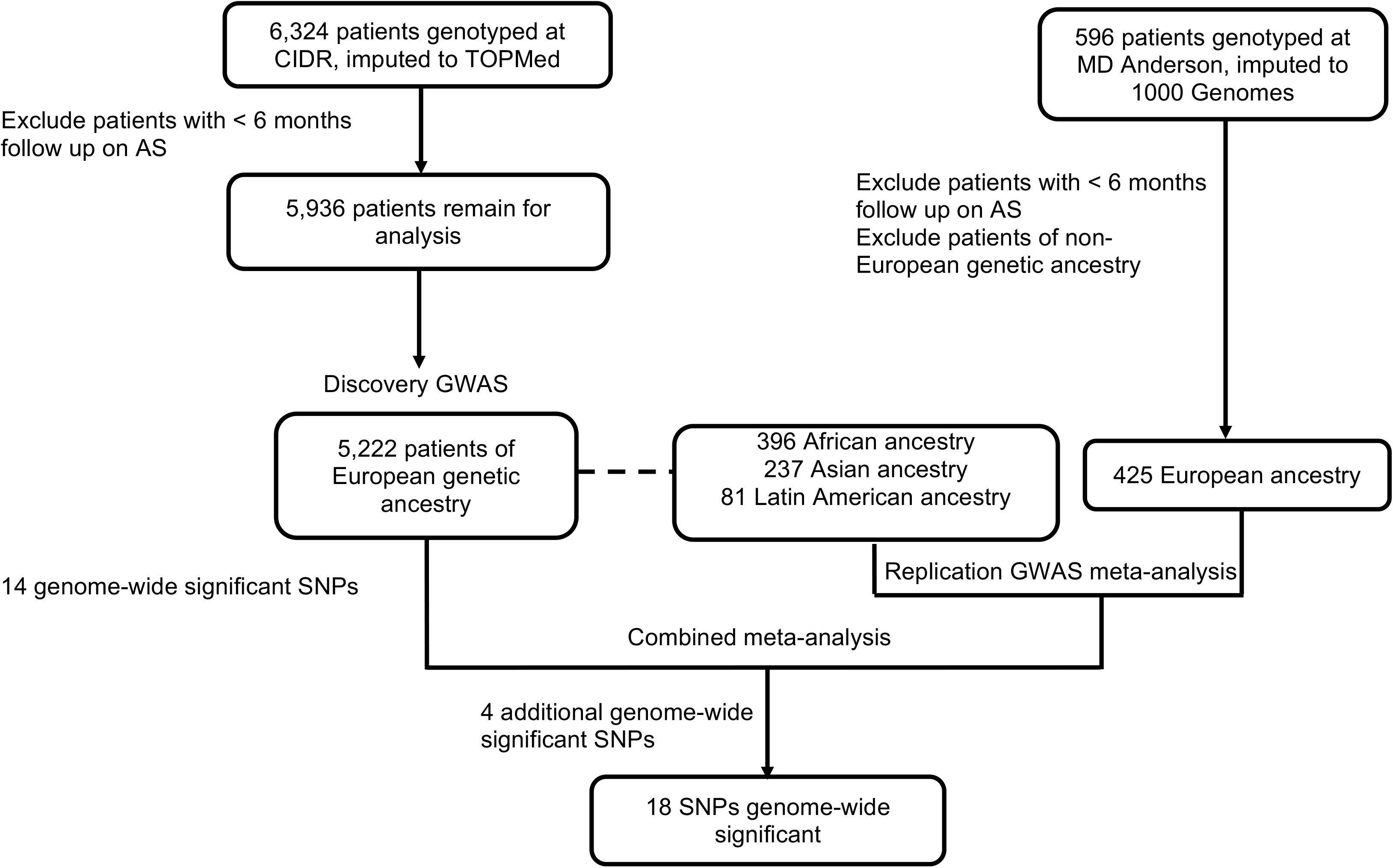
Flow chart highlighting the approach and samples used in the genome-wide association analysis. First, we undertook a discovery GWAS in men of European ancestry. Fourteen SNPs were associated with conversion (*P* < 5 × 10^-8^). All SNPs were evaluated for replication in the replication cohorts alone and then in a meta-analysis combining the discovery and replication cohorts. Four additional SNPs reached statistical significance in the combined meta-analysis (*P* < 5 × 10^-8^).

**Figure 2.**
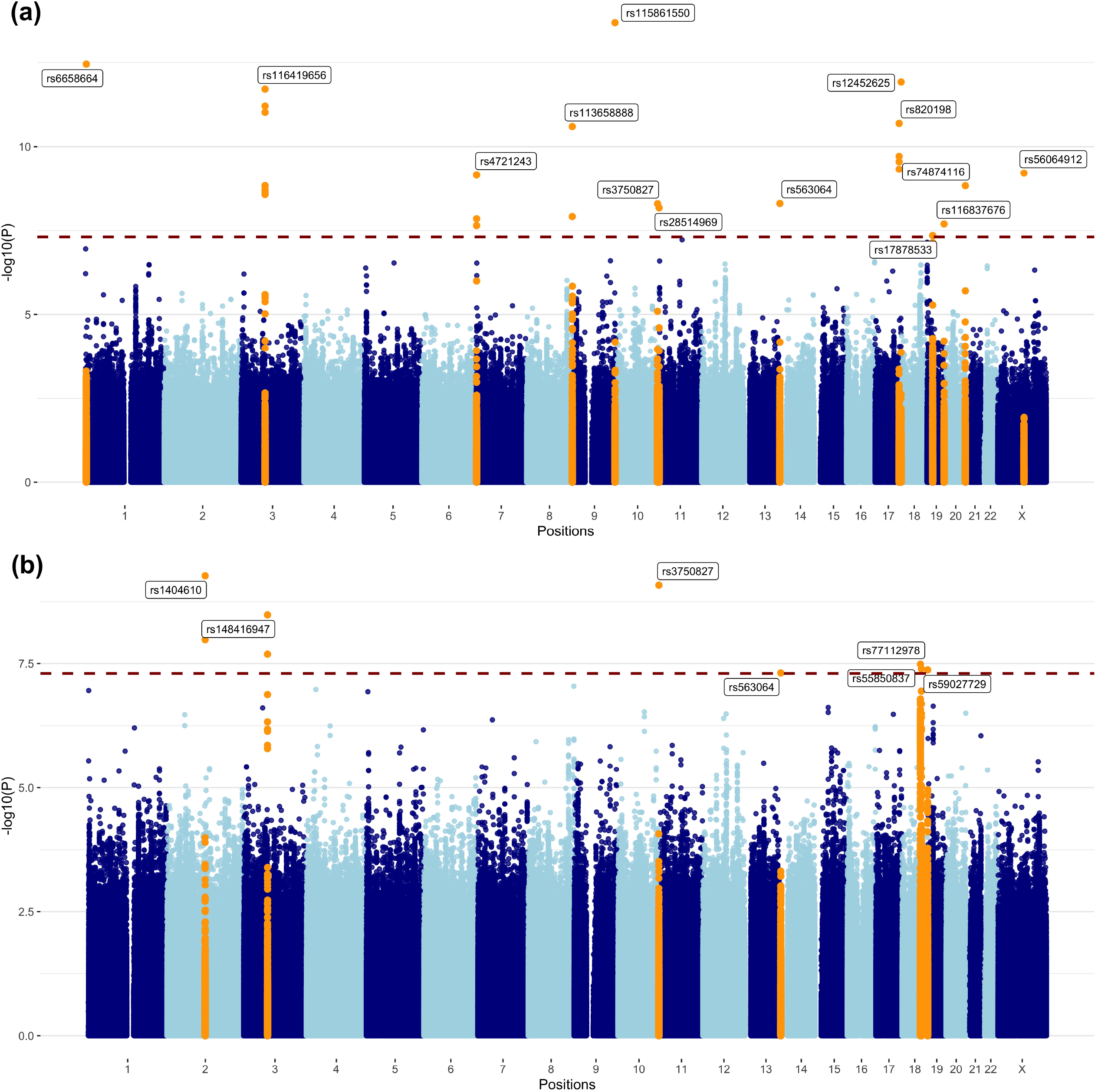
Results from the GWASs of conversion from AS to treatment. (a) in 5,222 prostate cancer patients of European ancestry; (b) in discovery and replication cohorts. P values are for variant associations with conversion, adjusted for age and ten ancestry principal components using Cox proportional hazards models. Blue dashed line denotes the genome-wide significance threshold. Orange peaks indicate genome-wide significant hits (*P* < 5 × 10^-8^). The top variants in each chromosome are annotated with their rsID.

Of the 18 SNPs, four were common (MAF > 0.01) and the remainder were low frequency (MAF = 0.01) (**Table 1**). Three were located within 1 Mb of previous PC GWAS-identified variants, although they were not in linkage disequilibrium with these variants (*r*^2^ < 0.3). These were: intronic SNP rs4721243 of *MAD1L1* (MIM: 602686) on chromosome 7 (CIDR European GWAS *HR* = 5.65, *P* = 7 × 10^-10^); rs1404610 near *GLI2* (MIM: 165230; combined meta-analysis *HR* = 3.74, *P* = 5.4 × 10^-10^) on chromosome 2; and rs74874116 near *GATA5* (MIM: 611496; CIDR European *HR* = 2.67, *P* = 1.47 × 10^-9^) on chromosome 20.

**Table 1.**
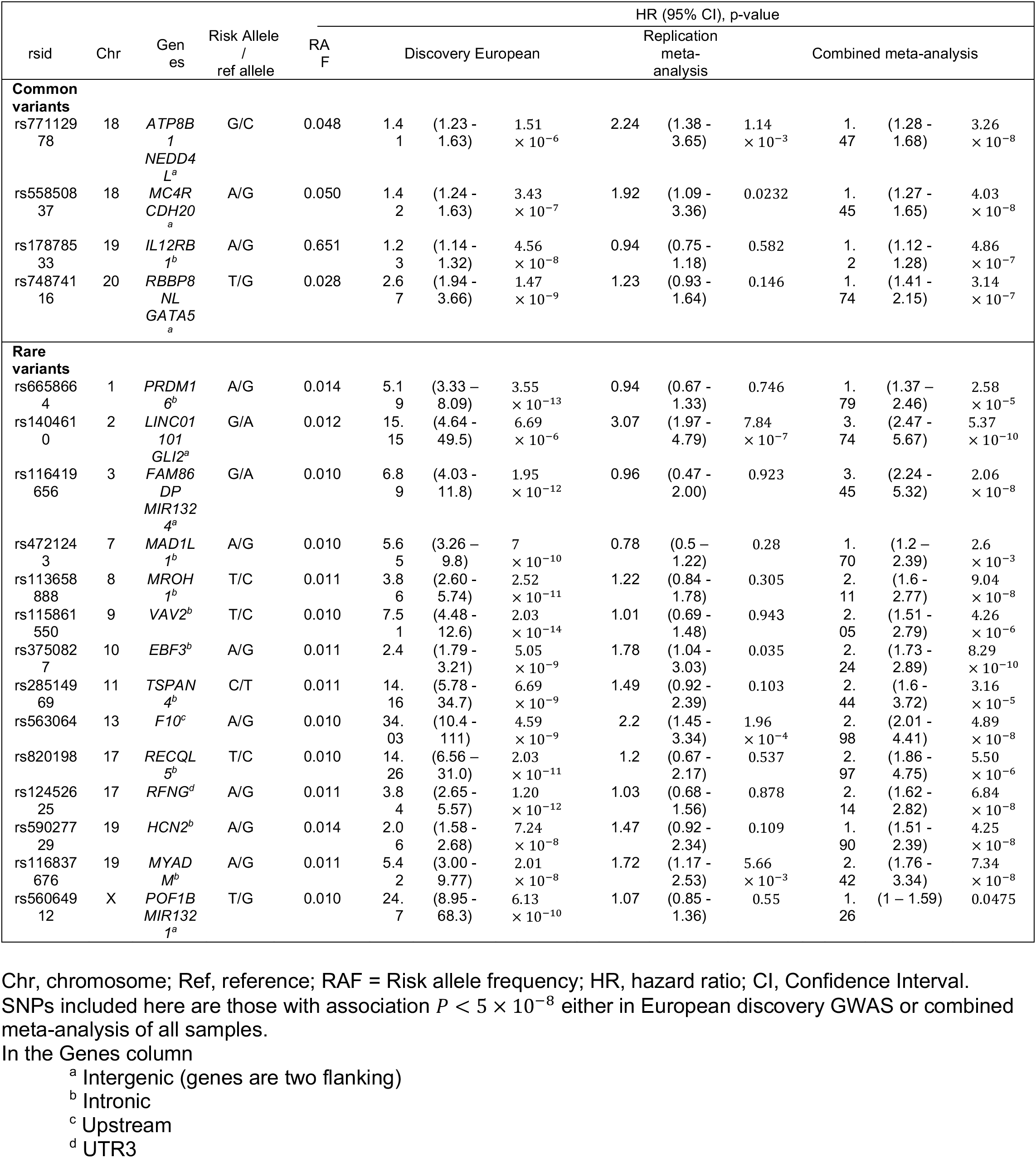
Results for 4 common and 14 variants associated with conversion from AS to treatment in a genome-wide association analysis.

### Transcriptome-Wide Association Study of Conversion from AS to Treatment

In the multi-tissue TWAS analysis using S-MultiXcan, the imputed expression levels of two genes were associated with conversion after Bonferroni correction for multiple testing: *MAST3* (MIM: 612258; P-value=6.9 × 10^-7^) and *GAB2* (MIM: 606203; P-value=2.0 × 10^-6^). Imputed expression levels of two other genes suggested association with conversion: *ARRDC2* (P-value=2.7 × 10^’,^) and *CELSR1* (MIM: 604523; P-value=9.5 × 10^-5^). When looking only at prostate tissue, we observed modest associations for *MAST3* (P-value=0.08) and *GAB2* (P-value=4.1 × 10^-5^), as well as a suggestive association between imputed expression of the gene *ZNF644* (MIM: 614159) and conversion (P-value=9.9 × 10^-5^).

### Genetic Risk Scores and Conversion from AS to Treatment

Increasing GRS for PC susceptibility (GRS_PC_) was positively associated with conversion from AS to treatment, even after adjusting for clinical covariates (Figure 3a; **Table S2**). The fully adjusted HR for conversion for men in the top decile of the GRS_PC_ compared to the middle two deciles was 1.13 (95% CI, 0.94-1.36; Figure 3a; **Table S2**). Men in the bottom 10^th^ percentile of the GRS_PC_ distribution had a significantly lower conversion rate than the middle two deciles of the GRS_PC_ (HR=0.69; 95% CI, 0.56-0.86; Figure 3a; **Table S2**).

**Figure 3.**
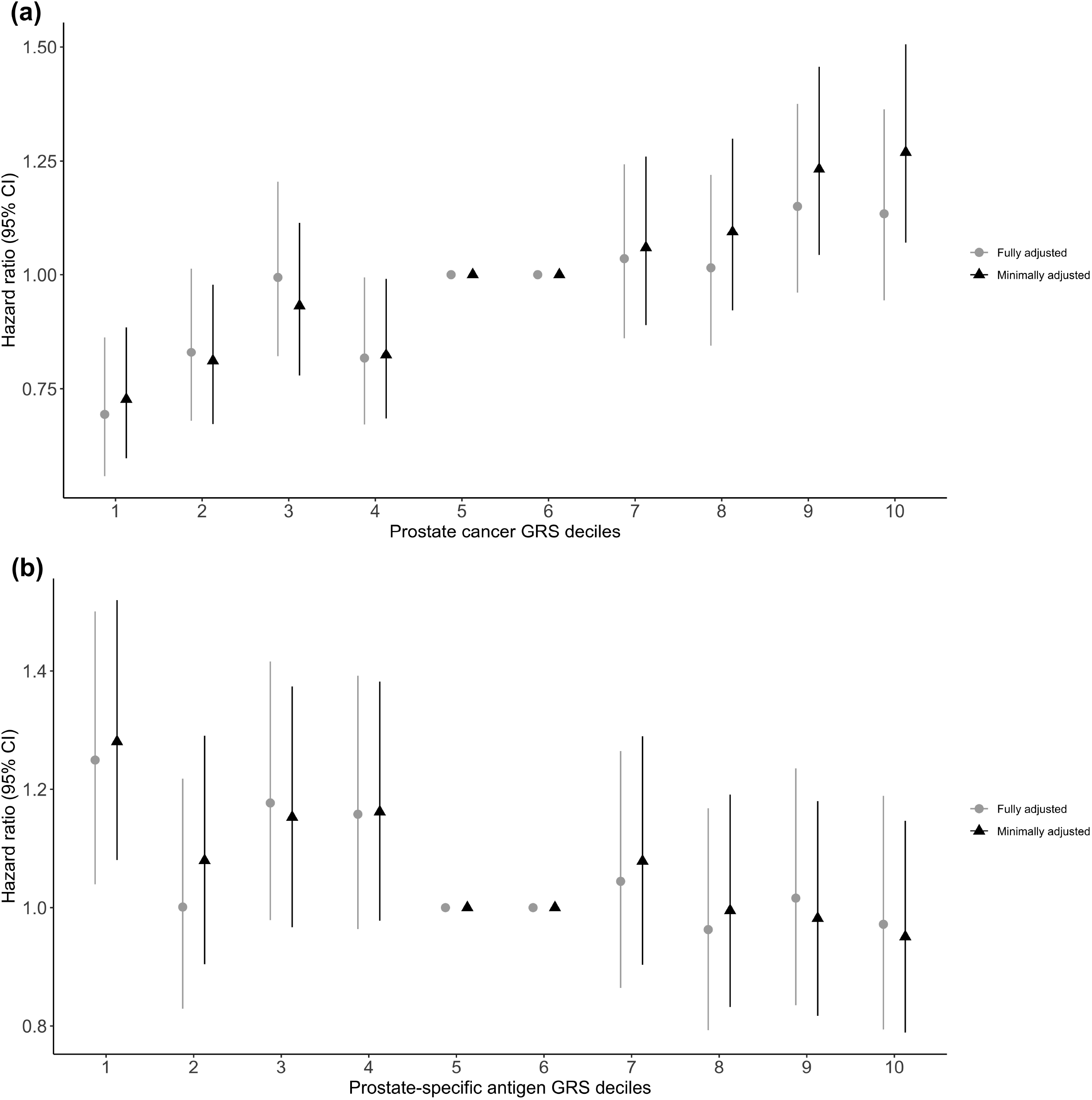
Association between time to conversion from AS to treatment. (a) with the prostate cancer GRS; (b) with the prostate-specific antigen GRS. The fifth and sixth deciles of prostate cancer GRS are used as the reference. Bars indicate 95% confidence intervals (CI) around the hazard ratio (HR) estimates. The minimally adjusted model includes age and the first 10 genetic prinicpal components. The fully adjusted model also includes Gleason grade group (GG1, GG2, or ≥ GG3); PSA concentration (ng/mL); clinical stage (cT1, cT2, or cT3/cT4); and number of positive biopsy cores (1-2, 3, or ≥ 4).

From the 36-variant GRS for PSA concentration (GRS_PSA_), we observed the opposite pattern: increasing GRS_PSA_ was inversely associated with conversion (Figure 3b; **Table S3**). Compared to the 40-60^th^ percentiles, men in the bottom 10^th^ percentile of the PSA GRS distribution experienced a shorter time to conversion (fully adjusted HR=1.25; 95% CI, 1.04-1.50; Figure 3b; **Table S3**).

### Potential Clinical Utility of the GRS

The time to conversion in the low- and intermediate-risk groups varied depending on whether men were in the top or bottom deciles of the GRS_PC_ and GRS_PSA_ distributions (Figure 4). For GRS_PC_, the Kaplan-Meier curves contrasting the top *versus* bottom deciles were significantly different for the low- and intermediate-risk groups (p=3×10^-5^ and p=0.016, respectively). Similarly, the top and bottom deciles of the GRS_PSA_ differed for the intermediate-risk groups (p=0.003). There was no clear difference between the deciles of GRS_PC_ or GRS_PSA_ in patients with high-risk disease.

**Figure 4.**
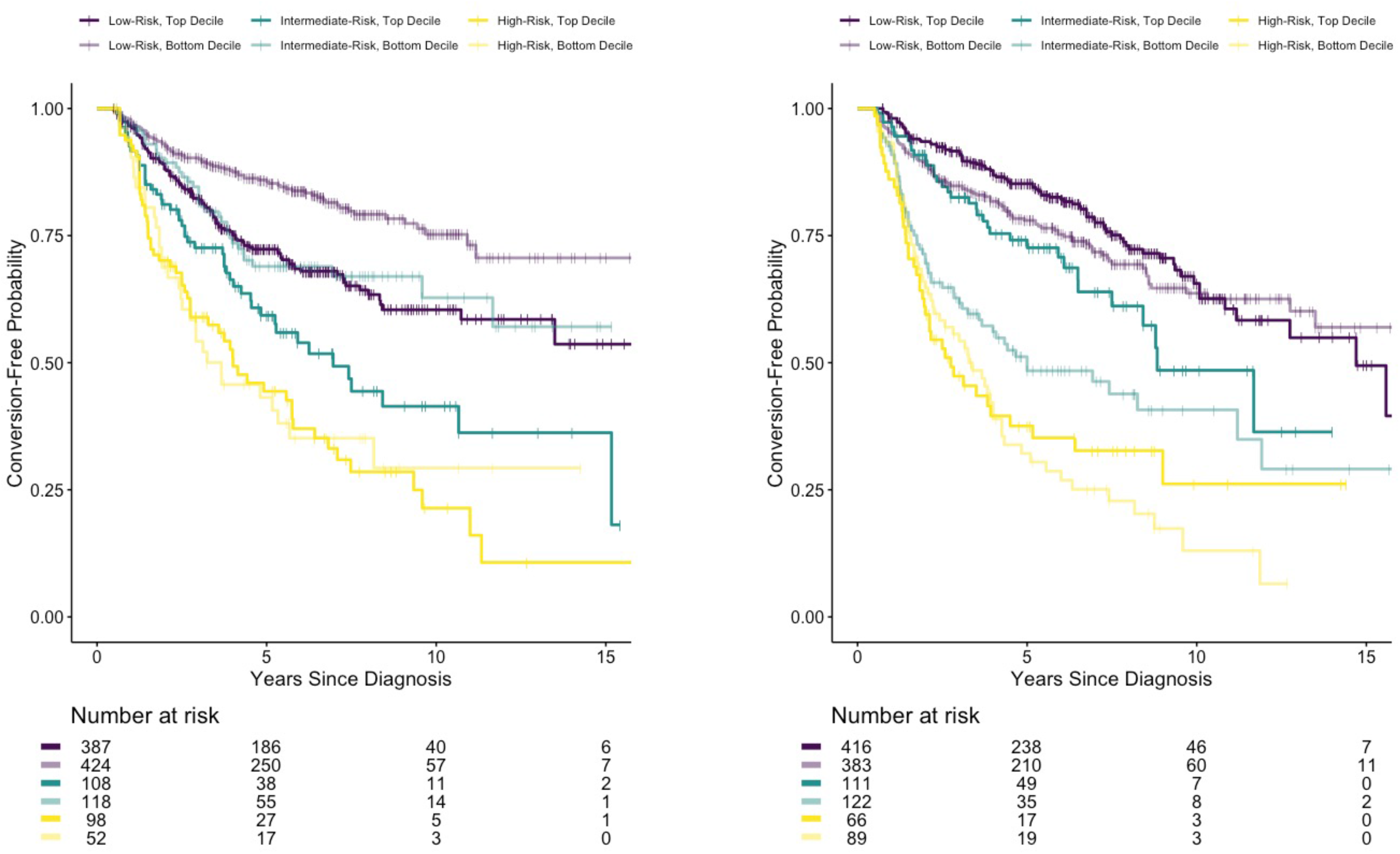
Kaplan-Meier plots of active surveillance conversion-free probability for low, intermediate, and high clinicopathological risk categories. The plots are stratified by the top and bottom deciles of genetic risk scores for prostate cancer (GRS_PC_, panel a) and for PSA levels (GRS_PSA_, panel b). The curves within each risk category are compared between the top and bottom GRS deciles using a log-rank test (P-values given next to corresponding curves).

A baseline model including age at diagnosis and principal components achieved an AUC = 0.55 for time to conversion. This was substantially improved by incorporating the clinical characteristics into the model: AUC = 0.653 (**Table S4**). Adding the GRS_PC_ to this model resulted in modest improvement (AUC=0.659). Augmenting this model with GRS_PSA_ produced minimal improvement (AUC = 0.661).

## Discussion

In this GWAS of PC patients managed with AS, we detected 18 novel SNPs and two candidate genes associated with conversion. We further found that GRS for PC susceptibility in addition to PSA level were associated with conversion, providing information beyond conventional clinical and pathologic measures of the disease. These findings provide preliminary support for using germline genetic information to inform the initial management of men with newly diagnosed, clinically localized PC.

Of the 18 SNPs associated with conversion, 15 were not previously associated with PC risk. These include a low-frequency (MAF = 0.01) intronic variant, rs4721243 at *MAD1L1*, at a previously identified PC locus^43^. The SNP was uncorrelated with the previously reported genome-wide significant PC SNP at the locus (rs4513875, *r*^2^ = 0.012 in the 1000 Genomes global reference data). One detected SNP (rs74874116) was 32kb away from a PC-associated indel (rs139135938), with little correlation (*r*^2^ = 0.015 in 1000 Genomes). The neighboring gene, *GATA5*, encodes a transcription factor that contains two GATA-type zinc fingers and is required during cardiovascular development^62^. This gene contains two variants previously associated with benign prostatic hyperplasia (MIM: 600082) and associated lower urinary tract symptoms^63^ (MIM: 618612). Another SNP in a PC risk locus was rs1404610, nearby *GLI2*, a transcription factor that one study found regulates the growth and tumorigenicity of prostate cells^64^.

Many of the novel SNPs we found to be associated with conversion are intronic, including SNPs in genes involved in cellular signaling, growth, and differentiation. *PRDM16* (MIM: 605557), where rs6658664 is located, is associated with evasion of apoptosis by prostatic cancer cells^65^. Intronic SNP rs115861550 in *VAV2* (MIM: 600428) is upregulated in human PC tumors and is a prognostic indicator for poor outcome^66^. Another intronic SNP, *EBF3* (MIM: 607407), has been shown to regulate the expression of genes involved in cell growth, proliferation, and apoptosis^67^. *RECQL5* (MIM: 603781), where SNP rs820198 is located, regulates DNA repair intermediate structures, and studies have observed elevated *RECQL5* expression in other cancers such as breast (MIM: 114480) and bladder (MIM: 109800)^68–70^. SNP rs820198 is annotated to an active CTCF (CCCTC-binding factor) binding site, and CTCF expression in linked to poor outcome in prostate cancer^71^. Although intergenic, SNP rs77112978 is near *NEDD4L* (MIM: 606384), whose expression is decreased in PC^72^. Intergenic SNP rs55850837-A, associated with conversion in our study, was associated with reduced body mass index^73^ and body fat percentage^74^ in the phenome-wide association data curated by the IEU OpenGWAS Project^75^. SNP rs12452625, a UTR’3 variant of *RFNG* (MIM: 602578) gene, is correlated with variants associated with multiple traits, including heel bone mineral density, lung function, and waist-hip ratio^76, 77^. This SNP is also predicted to be a functional target of microRNA hsa-miR-629-3p, which may serve as a biomarker for lung metastases of triple-negative breast cancer^78^.

Our TWAS suggests a possible role for *MAST3* and *GAB2* in conversion. A study described *MAST3* as an inflammatory bowel disease (IBD) susceptibility gene that regulates NF-κB activity through TLR4^79^. Two recent studies have described increased risk for PC in men with IBD (MIM: 601458)^80, 81^. Regarding *GAB2,* the knockdown of this gene in PC cells altered the expression of over 1,200 genes and inhibited p53 signaling^82^.

We found that the PC GRS based on 269 known risk variants was positively associated with conversion. Moreover, a PSA GRS based on 36 known genetic variants for PSA levels exhibited a weak inverse association with conversion. We expected these GRS to have opposite directions of effect on conversion, given that the PSA GRS may reflect the potential ascertainment of higher-risk PC in men with lower genetically predicted PSA levels. While the overall GRS only contributed modest model discrimination beyond established risk factors for conversion, the associations observed in the tails (i.e., deciles) of the GRS distribution were most pronounced among men in low and intermediate clinicopathological categories. This finding suggests that men with lower-risk disease, but high PC GRS (or low PSA GRS) may be more likely candidates for early treatment or possibly a higher intensity of surveillance. A recent study of European ancestry men with low-risk PC managed on AS reported associations between higher PC GRS with more positive cores and with bilateral tumor location at diagnostic and surveillance biopsy^83^; note that ∼50% of the men in this previous study are also included here, comprising ∼10% of our study population.

Strengths of this study include leveraging a large, multi-institutional collaborative study of AS to model the effects of genetic risk variants independent of clinical risk parameters. Sixty-three percent of the replication sample (n = 714) were men of non-European genetic ancestry, allowing us to test the generalizability of the SNPs discovered in the European sample. Our GRS included the most recently available GWAS weights from PC and PSA. Limitations of our study included the lack of confirmatory or surveillance biopsies to reduce misclassification of clinical parameters at diagnosis and follow-up. In addition, conversion could conflate disease progression with patient anxiety and/or physician preference for AS management. However, discontinuing AS due to anxiety was relatively uncommon in this study (about 6% of events), and these cases were censored in the GWAS analysis. Furthermore, given the relatively short follow-up for more robust PC outcomes, the sample sizes for PSA failure after treatment (n=124), metastases (n=29), or PC-specific death (n=11) are too small for a GWAS analysis.

In summary, we have undertaken the first GWAS of conversion among men diagnosed with PC. This multi-institutional study detected a genetic basis of conversion, suggesting that genetic factors may provide valuable information to stratify men with PC by their risk of discontinuing AS. Important future work will expand this study to more men placed on AS, increasing our ability to detect genetic variants associated with conversion. This may in turn help address concerns that biopsy sampling may underestimate a tumor’s aggressiveness and provide a more personalized approach to decisions surrounding AS.

## Data Availability

The MEGA data analyzed in this publication have been deposited in dbGap and are accessible through dbGap Study Accession number phs002056.v1.p1.

## Supplemental Data

Supplemental data include one figure and four tables.

## Declarations

This study was approved by the Robert H. Lurie Comprehensive Cancer Center of Northwestern University Scientific Review (IRB) Committees. The approval number is STU00077147, which was most recently given annual approval on 7/8/2021.

## Acknowledgements

**Funding/Support:** P50CA180995 (William J. Catalona) 08/01/15 – 07/31/20 NIH/NCI SPORE in Prostate Cancer PI on the SPORE grant, Center for Inherited Disease Research (CIDR) award from the NCI for patient sample genotyping (X01HG009642), Urological Research Foundation (William J. Catalona), NorthShore University Health System, Evanston, IL, USA (Brian T. Helfand). Additional support was provided by award numbers R01CA158627 and R01CA195505 (Leonard S. Marks), P50 CA186786 (Todd M. Morgan), UL1 TR000445 from NCATS/NIH for Vanderbilt REDCap (Daniel A. Barocas), P50 CA097186 and K05 CA175147 (Janet L. Stanford), and U01 CA113913 (Martin G. Sanda) from the National Institute of Health, and W81XWH-13-2-0074 (Peter R. Carroll) from the Department of Defense.

## Web Resources

METAL (https://genome.sph.umich.edu/wiki/METAL_Documentation)

PLINK v1.9 (https://www.cog-genomics.org/plink/)

MetaXcan, S-MetaXcan (https://github.com/hakyimlab/MetaXcan)

IEU OpenGWAS Project (https://gwas.mrcieu.ac.uk/)

Online Mendelian Inheritance in Man (http://www.omim.org)

