## Supplemental data for "Genetic Factors Associated with Prostate Cancer Conversion from Active Surveillance to Treatment"

**Supplemental Information**

**Figure S1. QQ plots of the GWASs.** (a) QQ plot of GWAS in individuals of European ancestry. (b) QQ plot of combined GWAS meta-analysis of all individuals.

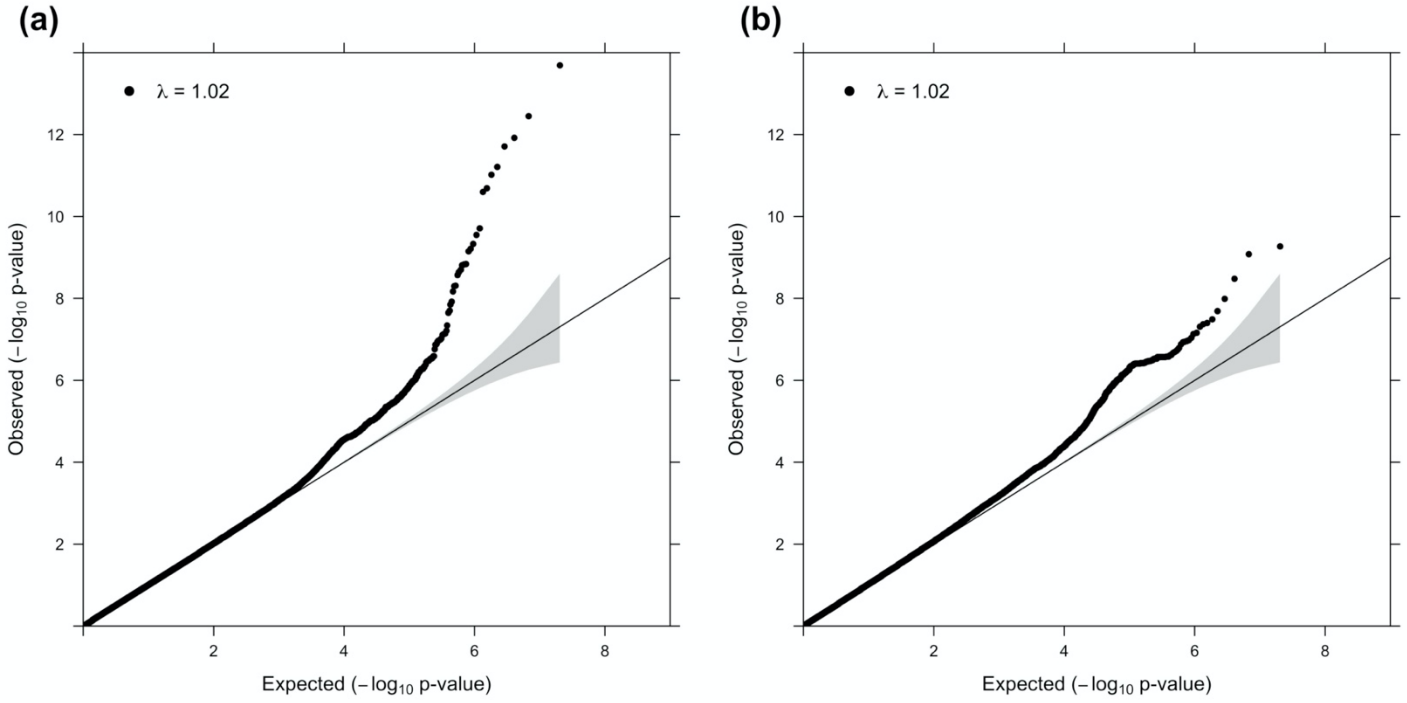

**Table S1. Characteristics of active surveillance patients included in the discovery and replication genome-wide association study.**

| | Discovery | | Replication | | | | | | | |
|---|---|---|---|---|---|---|---|---|---|---|
| Ancestral population | European | | African | | Asian | | Latin American | | European[a] | |
| Sample size | 5,222 | | 396 | | 237 | | 81 | | 425 | |
| Mean age (± s.d.) | 63.5 | (±7.2) | 62 | (±8.2) | 63.8 | (±7.7) | 63 | (±7.2) | 64.4 | (±8.2) |
| Median follow-up time (months, IQR) | 80 | (51,112) | 68 | (40,106) | 86 | (54,118) | 59 | (33,85) | 80 | (31,110) |
| Gleason grade | | | | | | | | | | |
| GG1 | 4,819 | (92%) | 359 | (91%) | 219 | (92%) | 77 | (95%) | 372 | (87%) |
| GG2 | 344 | (7%) | 32 | (8%) | 15 | (6%) | 4 | (5%) | 48 | (11%) |
| ≥ GG3 | 55 | (1%) | 4 | (1%) | 3 | (1%) | 0 | (0%) | 5 | (1%) |
| PSA (MAD) | 5.0 | (±2.1) | 5.3 | (±2.0) | 5.3 | (±2.5) | 4.7 | (±2.4) | 4.1 | (±2.2) |
| Clinical tumor stage | | | | | | | | | | |
| cT1 | 4,138 | 79% | 322 | 81% | 161 | 68% | 66 | 81% | 372 | 88% |
| cT2 | 641 | 12% | 49 | 12% | 39 | 16% | 9 | 11% | 53 | 12% |
| cT3 or cT4 | 34 | 1% | 5 | 1% | 5 | 2% | 1 | 1% | 0 | 0% |
| Number of cores | | | | | | | | | | |
| 1-2 | 4,113 | (79%) | 298 | (75%) | 177 | (75%) | 67 | (83%) | 386 | (91%) |
| 3 | 451 | (9%) | 44 | (11%) | 20 | (8%) | 7 | (9%) | 27 | (6%) |
| ≥ 4 | 522 | (10%) | 39 | (10%) | 37 | (16%) | 7 | (9%) | 12 | (3%) |
| PC risk category | | | | | | | | | | |
| Low | 3,639 | (70%) | 262 | (66%) | 141 | (59%) | 59 | (73%) | 314 | (74%) |
| Intermediate | 983 | (19%) | 82 | (21%) | 53 | (22%) | 14 | (17%) | 91 | (21%) |
| High | 599 | (11%) | 52 | (13%) | 43 | (18%) | 8 | (10%) | 20 | (5%) |

a. MD Anderson samples of European Ancestry.
IQR, interquartile range; MAD, median absolute deviation; PSA, prostate-specific antigen; PC, prostate cancer.
Men of genetically inferred European ancestry genotyped by CIDR are included in the discovery GWAS. The other participants genotyped by CIDR and men from MD Anderson are included in the replication.
Age and cancer clinical characteristics were measured at diagnosis.
PC risk categories:

Low-risk patients had all the following criteria: GG1 only (Gleason ≤ 3+3), PSA <10 ng/mL, clinical stage T1, and ≤ 3 positive biopsy cores.

Intermediate-risk patients had any of the following, with no high-risk criteria: GG2 (Gleason 3+4), PSA 10-20 ng/mL, or clinical stage T2.

High-risk patients had any of the following: ≥ GG3 (≥ Gleason 4+3), PSA ≥20 ng/mL, clinical stage ≥T3, or ≥ 4 positive biopsy cores.

Percentages do not all sum to 100% due to missingness.

**Table S2. Hazard ratios for the association between time to AS failure with prostate cancer GRS.**

| | Minimally Adjusted | | | Fully Adjusted | | |
|---|---|---|---|---|---|---|
| | HR[a] | 95% CI | P | HR[b] | 95% CI | P |
| **Decile** | | | | | | |
| 0-10 | 0.73 | 0.60, 0.89 | 0.0016 | 0.69 | 0.56, 0.86 | 0.0010 |
| 10-20 | 0.81 | 0.68, 0.98 | 0.030 | 0.83 | 0.68, 1.01 | 0.067 |
| 20-30 | 0.93 | 0.78, 1.11 | 0.44 | 0.99 | 0.82, 1.20 | 0.95 |
| 30-40 | 0.83 | 0.69, 0.99 | 0.041 | 0.82 | 0.67, 0.99 | 0.043 |
| 40-60 | 1.00 | Reference | | 1.00 | Reference | |
| 60-70 | 1.06 | 0.89, 1.26 | 0.50 | 1.03 | 0.86, 1.24 | 0.72 |
| 70-80 | 1.10 | 0.92, 1.30 | 0.30 | 1.02 | 0.85, 1.22 | 0.87 |
| 80-90 | 1.23 | 1.04, 1.46 | 0.014 | 1.15 | 0.96, 1.37 | 0.13 |
| 90-100 | 1.27 | 1.07, 1.51 | 0.0061 | 1.13 | 0.94, 1.36 | 0.18 |

HR = Hazard Ratio; CI = Confidence Interval

[a] Hazard ratios are adjusted for age and the first 10 genetic principal components

[b] Hazard ratios are adjusted for age, the first 10 genetic principal components, Gleason grade group (GG1, GG2, or ≥ GG3); PSA concentration (ng/mL); clinical stage (cT1, cT2, or cT3/cT4); and number of positive biopsy cores (1-2, 3, or ≥ 4).

**Table S3. Hazard ratios for the association between time to AS failure with prostate-specific antigen GRS.**

| | Minimally Adjusted | | | Fully Adjusted | | |
|---|---|---|---|---|---|---|
| | HR[a] | 95% CI | P | HR[b] | 95% CI | P |
| **Decile** | | | | | | |
| 0-10 | 1.28 | 1.08, 1.52 | 0.0044 | 1.25 | 1.04, 1.50 | 0.017 |
| 10-20 | 1.08 | 0.90, 1.29 | 0.40 | 1.01 | 0.83, 1.22 | 0.96 |
| 20-30 | 1.15 | 0.97, 1.37 | 0.11 | 1.18 | 0.98, 1.42 | 0.083 |
| 30-40 | 1.16 | 0.98, 1.38 | 0.088 | 1.16 | 0.96, 1.39 | 0.12 |
| 40-60 | 1.00 | Reference | | 1.00 | Reference | |
| 60-70 | 1.08 | 0.90, 1.29 | 0.40 | 1.05 | 0.86, 1.27 | 0.65 |
| 70-80 | 1.00 | 0.83, 1.19 | 0.96 | 0.96 | 0.79, 1.17 | 0.70 |
| 80-90 | 0.98 | 0.82, 1.18 | 0.84 | 1.02 | 0.84, 1.24 | 0.87 |
| 90-100 | 0.95 | 0.79, 1.15 | 0.60 | 0.97 | 0.79, 1.19 | 0.78 |

HR = Hazard Ratio; CI = Confidence Interval

[a] Hazard ratios are adjusted for age and the first 10 genetic principal components

[b] Hazard ratios are adjusted for age, the first 10 genetic principal components, Gleason grade group (GG1, GG2, or ≥ GG3); PSA concentration (ng/mL); clinical stage (cT1, cT2, or cT3/cT4); and number of positive biopsy cores (1-2, 3, or ≥ 4).

**Table S4**. **ROC analysis of PC and PSA GRS compared to clinical characteristics for conversion from AS to treatment.**

| Model | $GRS_{PC}$[d] | | | $GRS_{PSA}$[d] | | | AUC |
|---|---|---|---|---|---|---|---|
| | HR | 95% CI | P-value | HR | 95% CI | P-value | |
| Reference model[a] | | | | | | | 0.550 |
| PC clinical characteristics[b] | | | | | | | 0.653 |
| PC clinical characteristics + PC GRS[c] | 1.13 | 1.07, 1.19 | $3.3 \times 10^{-6}$ | | | | 0.659 |
| PC clinical characteristics + PSA GRS[c] | | | | 0.94 | 0.89, 0.98 | $8.5 \times 10^{-3}$ | 0.655 |
| PC clinical characteristics + PC GRS + PSA GRS | 1.15 | 1.09, 1.22 | $1.5 \times 10^{-7}$ | 0.91 | 0.87, 0.96 | $3.0 \times 10^{-4}$ | 0.661 |

[a] All models contain age and the first 10 principal components.
[b] PC clinical characteristics were Gleason grade group (GG1, GG2, or ≥ GG3); PSA concentration (ng/mL); clinical stage (cT1, cT2, or cT3/cT4); and number of positive biopsy cores (1-2, 3, or ≥ 4).
[c] PC GRS is constructed from 269 prostate cancer associated variants, while PSA GRS is derived from 36 PSA risk variants.
[d] The test statistics of $GRS_{PC}$ and $GRS_{PSA}$ are reported when the model contains them.
